# Detecting accelerated retinal decline in mental disorders through normative modeling

**DOI:** 10.1101/2024.06.11.24308654

**Authors:** Foivos Georgiadis, Nils Kallen, Giacomo Cecere, Finn Rabe, Victoria Edkins, Daniel Barthelmes, Amber Roguski, Daniel Smith, Matthias Kirschner, Philipp Homan

## Abstract

**Objective:** Several studies have found thinner retinal tissue in mental disorders compared to healthy controls. Because the retina is part of the human brain, this suggests that informative brain structure readouts can be obtained efficiently through retinal imaging. Instead of focusing on group-level case-control differences, we used normative modeling to estimate age-related decline of the human retina (and its expected variation) and compared it to the decline seen in schizophrenia (SZ), bipolar disorder (BD), and major depression (MDD). We hypothesized accelerated retinal decline in mental disorders compared to controls, with SZ being most affected, followed by BD, then MDD.

**Methods:** Using UK Biobank data, we estimated age-related retinal decline in healthy controls (HC, N = 56,545) for total macular thickness (including coronal subfields) and two sublayers (retinal nerve fiber layer; RNFL; and ganglion cell-inner plexiform layer; GC-IPL). We then compared the decline in SZ (N = 171), BD (N = 256), and MDD (N = 102) to the normative decline in HC.

**Results:** For HC, the pattern of age-related decline for total macular thickness, RNFL, and GC-IPL was curve-like rather than linear and more pronounced in males compared to females. For mental disorders, the decline-pattern was generally faster, driven by SZ and disorder-specific macular subfields. There was also an enrichment of individuals with extremely low (infranormal) values. These results were confirmed in robustness checks that ruled out unspecific confounders.

**Conclusion:** These findings suggest that mental disorders, particularly SZ, involve accelerated neurodegenerative decline that can be detected in the human retina.

## Introduction

The human retina is an extension of brain tissue that directly connects with the outside world^1^ and as such can be non-invasively imaged to provide structural readouts of the central nervous system. Mirroring deficits found in central gray matter^2–6^ the retina has been found to be thinner compared to healthy controls in neurologic ^7–9^ and psychiatric disorders, including schizophrenia (SZ), bipolar disorder (BD) and major depression (MDD)^10–19^. Yet previous studies have mostly focused on group-level case-control differences which may mask the biological heterogeneity often seen in mental disorders and even healthy controls^20,21^ For example, just as the brain^22^, the retina shows marked variability associated with age and sex already in the healthy population^23,24^. When this healthy (or normative) variability is estimated with reasonable precision, charts of normative variability, akin to growth charts known from pediatrics, can be obtained through which individual disease cases can be compared to the expected normative variability. In other words, this approach, also known as normative modeling ^22,25–28^, is well-suited to provide a more personalized and dimensionally accurate view of mental disorders^26,28–30^. Accordingly, normative modeling has been used to quantify normative and disease-related variability as well as accelerated brain-aging in large-scale structural MRI studies^31–34^.

Here, we estimated such normative variability for the macular region, the area with the highest density of neurons in the retina ^1^, using the largest OCT sample of healthy control individuals to date (N = 56,545) from the UK Biobank. Our aim was to quantify the expected macular thickness decline ^23^ for the typical UK Biobank age-range (i.e., mid to late age), including its coronal subfields and two of its cellular sublayers, the retinal nerve fiber layer (RNFL) and the ganglion cell-inner plexiform layer (GC-IPL). Equipped with such normative retinal decline charts, we then located each of N = 538 individuals who were diagnosed with one of three major mental disorders (SZ, N = 171; BD, N = 265; and MDD, N = 102) in these charts, and measured their respective deviation from the norm. We hypothesized that age-related retinal decline would be more pronounced in mental disorders, with a gradient from SZ to BD to MDD^19^.

## Methods

### Design, setting, and participants

This analysis used cross-sectional measurements of optical coherence tomography (OCT) from the UK Biobank database (Figure 1A). The OCT measurement procedure, including inclusion and exclusion criteria and quality control metrics have been previously described^35^. Briefly, optical coherence tomography images were acquired using a spectral domain optical coherence tomography device, with a raster scan protocol of 6×6mm area centered on the fovea, consisting of 128 B-scans each with 512 A-scans, completed in 3.7 seconds. Automated analysis of retinal thickness was performed using custom software developed by Topcon Advanced Biomedical Imaging Laboratory, which used dual-scale gradient information for rapid segmentation of nine intraretinal boundaries, processing the images in approximately 120 seconds each. Here, we focused on retinal measurements of total macular thickness (Figure 1B), including the segregated macular subfields in the coronal plane, as well as macular sublayers; the retinal nerve fiber layer (RNFL, Figure 1B) and ganglion cell-inner plexiform layer (GC-IPL, Figure 1B). To estimate normative retinal decline, we used these measurements from N = 56,545 healthy individuals from the UK Biobank who had complete OCT imaging data for all retinal metrics (54.8% female; see Table 1 for demographics), including image quality control measurements (data fields 28552, 28553: OCT image quality) and without diagnosed eye disorders (data field 6148). To relate disease-related retinal decline to the normative decline (Figure 1C), we then included N = 538 separate individuals from the UK Biobank with a mental disorder (Table 1), including SZ (N = 171), BD (N = 265), and MDD (N = 102), according to ICD-10 primary or secondary diagnoses (F20, F31 and F33 respectively). As a robustness check, and because of the high prevalence of depressive symptoms in the UK Biobank, we subsequently also included individuals with a history of depressive symptoms, based on self-report of ever having experienced depressive symptoms for more than a week (Code 100349, see Supplementary Results).

**Figure 1.**
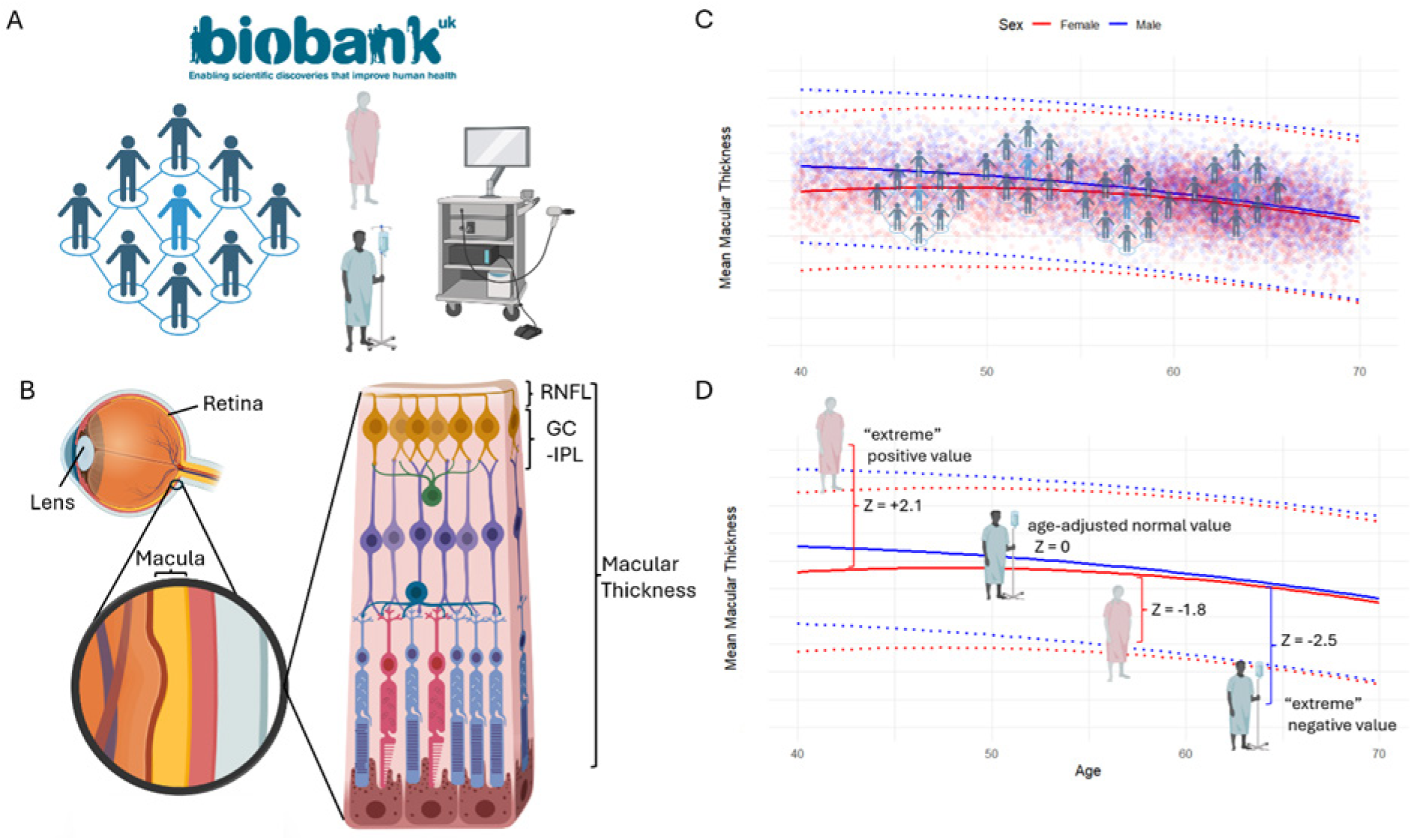
**A.** The UK Biobank has collected and standardized OCT measurements of more than 50 000 individuals providing an ideal resource for modeling the trajectory of retinal thickness across the mid-to late lifespan, as well as its variation; among the participating individuals, a small subset have been diagnosed with psychiatric disorders **B.** Left: Sagittal cross-section of the eye showing the localization of the lens and retina, including a magnification of the macula. In this study, we examine total macular thickness (macular thickness) as a single measurement, as well as segregated coronal subfields (see also Figure 3A); Right: schematic illustrations of the synaptic and cellular sublayers of the macula; in this study we also examine two outermost sublayers, namely the retinal nerve fiber layer (RNFL) and the combined ganglion cell and inner plexiform layers (GC-IPL). **C.** Normative curves of macular thickness across the mid-to late lifespan for female (red) and male (blue) individuals; solid lines represent the normative median (50^th^ centile), whereas dotted lines represent the 2.5^th^ (above) and 97.5^th^ (below) normative centiles. The individuals of the UK Biobank without ocular morbidities or a mental disorder diagnosis were used to develop normative models of retinal thickness (macular thickness, its coronal subfields, as well as the outermost sublayers: RNFL and GC-IPL). **D.** In a second step, patients with a diagnosis of SZ, BD or MDD were compared to their age- and sex-matched population norm, based on the normative curves; deviations from the age-adjusted normative values (solid lines) are quantified as z-scores, which are interpreted as the number of SDs each individual deviates from the norm. In a normal distribution, 95% of the population, have values between the 2.5^th^ and 97.5^th^ centile, or 1.96 SDs away from the norm. A z-score greater than 1.96 (in absolute value) is classified as an “extreme” deviation.

**Table 1.** Sample characteristics.

| Variable |  | HC |  | MD |  | BD |  | SZ |  |
| --- | --- | --- | --- | --- | --- | --- | --- | --- | --- |
|  |  | N | Mean (SD)/<br>Percentage | N | Mean (SD)/<br>Percentage | N | Mean (SD)/<br>Percentage | N | Mean (SD)/<br>Percentage |
| Age |  | 56545 | 56.26<br>(8.13) | 102 | 54.80<br>(8.28) | 265 | 54.83<br>(8.28) | 171 | 54.88<br>(8.70) |
| Sex: | Male | 25574 | 45.23 | 30 | 29.41 | 104 | 39.25 | 100 | 58.48 |
|  | Female | 30971 | 54.77 | 72 | 70.59 | 161 | 60.75 | 71 | 41.52 |
| Socioeconomic Status |  | 56545 | -1.10<br>(2.95) | 102 | 0.16<br>(3.57) | 265 | -0.26<br>(3.39) | 171 | 1.63<br>(3.60) |
| Body Mass Index |  | 56545 | 27.25<br>(4.72) | 102 | 27.82<br>(5.85) | 265 | 28.45<br>(5.26) | 171 | 28.47<br>(5.72) |
| Macular Thickness |  | 56545 | 277.14<br>(14.09) | 102 | 275.96<br>(13.94) | 265 | 274.30<br>(14.72) | 171 | 271.29<br>(16.53) |
| Retinal Nerve Fiber Layer Thickness |  | 56545 | 28.12<br>(4.81) | 102 | 28.76<br>(5.02) | 265 | 27.88<br>(4.73) | 171 | 27.86<br>(4.46) |
| Ganglion Cell-Intersitial Plexus Layer Thickness |  | 56545 | 73.84<br>(6.14) | 102 | 73.33<br>(6.08) | 265 | 73.11<br>(6.42) | 171 | 72.33<br>(6.34) |
| Image Quality |  | 56545 | 65.46<br>(11.57) | 102 | 67.13<br>(9.57) | 265 | 65.89<br>(10.75) | 171 | 65.15<br>(12.39) |
| Alcohol | Current | 52105 | 92.15 | 90 | 88.24 | 233 | 87.92 | 145 | 84.8 |
|  | Never | 2526 | 4.47 | 3 | 2.94 | 12 | 4.53 | 11 | 6.43 |
|  | Prefer not to answer | 46 | 0.08 | 2 | 1.96 | 1 | 0.38 | 1 | 0.58 |
|  | Previous | 1868 | 3.3 | 7 | 6.86 | 19 | 7.17 | 14 | 8.19 |
| DML | FALSE | 55296 | 97.79 | 100 | 98.04 | 261 | 98.49 | 163 | 95.32 |
|  | TRUE | 1249 | 2.21 | 2 | 1.96 | 4 | 1.51 | 8 | 4.68 |
| Ethnicity | African | 648 | 1.15 | 1 | 0.98 | 1 | 0.38 | 6 | 3.51 |
|  | Any other Asian | 334 | 0.59 | 6 | 5.88 | 2 | 0.75 | 1 | 0.58 |
|  | Any other mixed | 162 | 0.29 | 85 | 83.33 | 17 | 6.42 | 3 | 1.75 |
|  | Any other white | 2556 | 4.52 | 5 | 4.9 | 216 | 81.51 | 7 | 4.09 |
|  | British | 47445 | 83.91 | 4 | 3.92 | 4 | 1.51 | 130 | 76.02 |
|  | Caribbean | 1028 | 1.82 | 1 | 0.98 | 4 | 1.51 | 8 | 4.68 |
|  | Chinese | 242 | 0.43 | NA | NA | 15 | 5.66 | 1 | 0.58 |
|  | Indian | 1096 | 1.94 | NA | NA | 4 | 1.51 | 1 | 0.58 |
|  | Irish | 1895 | 3.35 | NA | NA | 1 | 0.38 | 10 | 5.85 |
|  | Other ethnic group | 798 | 1.41 | NA | NA | 1 | 0.38 | 2 | 1.17 |
|  | Pakistani | 195 | 0.34 | NA | NA | NA | NA | 1 | 0.58 |
|  | White and Asian | 146 | 0.26 | NA | NA | NA | NA | 1 | 0.58 |
| Smoke | Current | 5335 | 9.43 | 22 | 21.57 | 49 | 18.49 | 43 | 25.15 |
|  | Never | 31622 | 55.92 | 39 | 38.24 | 119 | 44.91 | 72 | 42.11 |
|  | Prefer not to answer | 168 | 0.3 | 41 | 40.2 | 97 | 36.6 | 2 | 1.17 |
|  | Previous | 19420 | 34.34 | NA | NA | NA | NA | 54 | 31.58 |

### Outlier removal

Similar to previous work, to arrive at our final study sample, outlier values were excluded using an interquartile range (IQR) trimming approach^30^. This approach removes observations above or below N*IQRs from the median, which are considered unreliable measurements (e.g. 1.5*IQRs, removes ∼5% of individuals, assuming a normally distributed variable). A lower value for the IQR multiplier excludes more participants than a higher value. To avoid overly strict exclusions, we repeated our analysis across IQR values to show the stability of our results (see Supplementary Figure S1). We report the result values for an IQR of 4, which under the assumption of a normally distributed variable only removes 0.01% of observations. Outlier removal was performed separately within the control population and each diagnostic group, to ensure that possible disease-related deviations from the population norm would not be removed as outliers. Lastly, to exclude the possibility that increased variability of patients’ retinal measurements (as compared to controls) would result in an overrepresentation and thus exclusion of “false-positive” outliers in the patient group, an enrichment analysis (using chi-squared statistics) of outlier data points in each group (HC, MDD, BD, SZ) was performed which was non-significant for differences in outlier observation rate (all χ2 < 1.86, all p> 0.17). Hence, we did not find evidence that the four groups differed significantly in the proportion of outliers per group.

### Normative model generation and specification

#### Dependent variables

To model decline in macular thickness, its coronal subfields, as well as RNFL and GC-IPL, we used GAMLSS in R^36^, a flexible statistical framework modeling non-linear relationships between predictors and outcomes. We built sex- and laterality-specific models for each metric similar to previous work^22,30^. For all downstream analyses, such as the description of the normative curves, as well as the analyses on z-scores, z-scores from the left and right eye were averaged, according to literature standards ^22,30,37^ (Figure 2A). To ensure robustness, we repeated the comparison of each patient group to the norm with a mixed model accounting for repeated measures (Participant ID as a random effect) with the non-averaged z-scores (Supplementary Results). Interaction effects of laterality with age and sex were further explored in a separate mixed model analysis (Supplementary Results, Figure S2).

**Figure 2.**
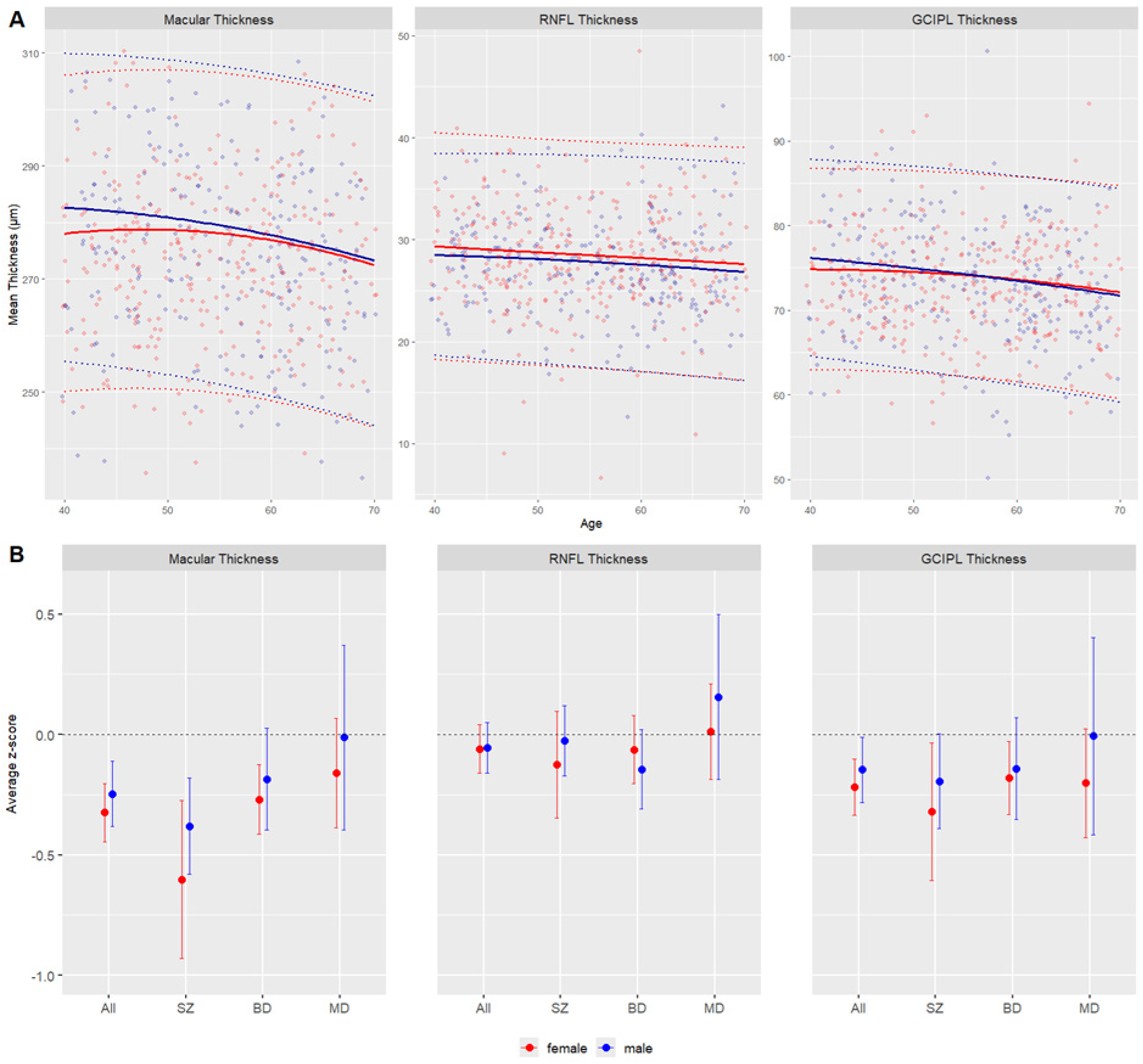
**A.** Normative age curves, estimated for healthy control females (red) and males (blue) and averaged across the left and right eye for macular thickness, retinal nerve fiber layer (RNFL) thickness and ganglion cell-inner plexiform layer (GC-IPL) thickness. The patient data points (raw values) are plotted on top of the normative curves. The normative mean for healthy females and males is shown in solid red and blue lines, respectively, with the extreme value boundaries (mean and 1.96 SD) indicated by dotted lines of the corresponding color. To enable a bivariate visualization, estimation of the mean (solid lines) and SD (dotted lines) of the HC assumed the mode and median values for ethnic background and image quality respectively **B.** Sex-stratified average deviations from the normative mean (z-score = 0), with error bars indicating 95% confidence intervals, for macular thickness, retinal nerve fiber layer thickness, and ganglion cell-inner plexiform layer thickness, for the combined transdiagnostic sample and for schizophrenia (SZ), bipolar disorder (BD), and major depressive disorder (MD) as separate groups. Error bars not touching the zero line indicate an average group z-score that is significantly different from zero.

#### Predictor variables and covariates

To model the effects of age on retinal thickness, which are known to be non-linear^23,38^, we used fractional polynomials which have been shown to provide an accurate method to model non-linear effects ^22,30^. Image quality and reported ethnic background were also included as covariates, since they have been shown to influence retinal parameters^38^. Although cardiovascular risk factors are also well known to have an effect on retinal thickness, most previous studies on retinal thickness deviations in patients with mental disorders did not have access to the phenotyping depth of the UKBB, and did not take them into account ^10–19^ Thus, we include cardiovascular risk factors and other covariates in our robustness analysis (see Supplementary Results), and not in our main analysis in order to be able to compare our main findings with the existing literature. Similar to previous work on retinal parameters in the UK Biobank^12^ we considered the following variables as cardiovascular risk factors, including body mass index (BMI), smoking status, alcohol drinker status, and an ICD-confirmed diagnosis of diabetes mellitus type 1 or 2. Finally, we also considered socioeconomic status (quantified by the Townsend Deprivation Index in the UK Biobank)^12^.

#### Distribution choice

The GAMLSS approach allows modelling of the entire statistical distribution of phenotype of a variable of interest across the predictor variables^22,27,36^. This means that the model not only predicts the mean (μ) across the lifespan but can model changes in standard deviation (σ) and further distribution parameters such as kurtosis (ν) and skewness (τ). The normal distribution is described by μ and σ, whereas more complex distributions can model the four distribution characteristics with increasing complexity. To select the appropriate distribution for our model, we compared the performance of several distributions, quantified by mean squared error, in an 80-20 5-fold cross validation framework (Figure S2). The normal distribution showed equivalent performance to the more complex distributions and was therefore chosen, since it provides the most parsimonious model (see Supplementary Results), and since using the next best performing distributions (generalized gamma or Box-Cox t) provided similar results (not reported).

### Normative curve description

To interpret the non-linear sex by age interaction patterns which can be observed from our normative curves (Figure 2A), we compared the right and left tail of the age distribution (younger than 45 and older than 65 years), for descriptive purposes. We described the normative curves of macular thickness, RNFL and GC-IPL by using the raw measurements (averaged left and right) as dependent variables, and age, sex and their interaction as predictors, while adjusting for ethnicity and image quality. In our Supplementary Results, we additionally explored laterality with a more complex mixed model with each individual’s eyes as repeated measures (lmer in R^39^) which measured interaction effects between the extremes of the age range (<45 years and >65 years), sex and laterality (see Supplementary results, Figure S3).

### Individual deviation scores

Our normative models result in age-specific predictions for each retinal metric, adjusting for covariates. Individual deviation scores (z-scores) from these normative predictions were subsequently computed for all individuals using the actual value and predicted μ and σ of the normal distribution for each data point (*pNO, qNO* functions from *gamlss* in R). Subsequently, for each individual, the z-scores for left and right eye were averaged into a single z-score, following previous literature and as confirmed by our replication using lateralized measurements (Supplementary Results). If an individual in the dataset had only a unilateral measurement and thus a unilateral z-score, then this measurement was used (0.8%, N = 452).

### Statistical analysis

All statistical analyses were carried out with the R-software version 4.3.0. To assess the statistical significance of differences in z-scores (Δz) between patients and controls and between diagnostic groups, we used linear models (lm) with the respective retinal thickness metric (macular thickness including subfields, RNFL, GC-IPL) deviations as the dependent variable and diagnostic group and sex as predictor variables. The alpha was set at 0.05. We report nominal p-values.

#### Extreme value analysis

In the framework of normative modeling, the interest is placed not only on the average z-score, but also on the extreme values of the distribution^31^. Therefore, based on the z-scores, for each of the HC, SZ, BP, MDD as well as the combined patient group, we estimated the percentage of individuals above and below 1.96 standard deviations, representing extreme values (infranormal and supranormal variability). To assess the statistical significance of difference in rate of occurrence (percentages) of extreme values in each of the groups, we used chi-squared tests.

### Robustness/sensitivity analyses

#### Robustness against overfitting

To exclude the possibility that our normative model was overfitting the training data, we performed a permutation analysis. A random 80% sample of the HC population was chosen at each iteration to train the normative models for each metric. Thereafter, normative deviations were compared between the remaining 20% of the healthy control population and the patient population; this circumvents any influence of overfitting since the HC sample compared to the patients has not been used to train the model.

#### Agreement of normative modeling results with linear modeling

We additionally confirmed our findings using a permutation approach with the simple linear model over 250 permutations (see Supplementary Results). Because of the large discrepancy in sample size we chose a random HC sample, ensuring a 3:1 HC to patient ratio (1614 HC: 538 patients), ensuring the same subsample of HC was used across the 3 metrics. We included the same covariates as in our original analysis apart from reported ethnicity. This exclusion was justified by the downsizing of the HC group and the concomitant presence of very small groups which provided unreliable estimates for this predictor in the linear model. This last analysis was repeated only on individuals of reported British ethnic background with very similar results (not reported). Since the z-score outcomes are directly interpretable as effect sizes, for the linear model we report both the measured effect and the Cohen’s *d* effect size.

## Results

### Description of normative decline

As seen in Figure 2A, we observed age-related thickness decline for all 3 retinal metrics (macular thickness, RNFL and GC-IPL), with female individuals qualitatively showing an attenuated slope compared to males for macular thickness and GC-IPL, but not RNFL. In macular thickness and GC-IPL thickness, higher values were found, on average, in male individuals, although the curves seemed to converge in late life, as a consequence of the attenuated age-related thickness decline in female individuals described above. In contrast, RNFL thickness was higher in female individuals across the lifespan. To quantify the non-linear interaction patterns described above, we compared the right and left tail of the age bins of our sample, i.e. individuals younger than 45 years and older than 65 years, as well as the interaction of age group with biological sex. More complex interaction effects between laterality, age and sex and were also explored with a linear mixed model and are detailed in the Supplement Results (see description of laterality effects in normative retinal trajectories).

#### Macular thickness

Compared to individuals younger than 45 years, the older age group’s (65+ years) macula was on average 7.42 µm thinner (CI = [−8.11, −6.73] µm, p < 0.001). Female individuals’ macula was thinner compared to males, with an average difference of 3.65 µm (CI = [−4.36, −2.96] µm, p < 0.001). The interaction effect between sex and age group was significant (estimate = 2.69µm, CI = [1.75, 3.64] µm, p < 0.001) and suggests that the reduction in macular thickness with age was less pronounced in females with female individuals in the older age group experienced a 2.69µm smaller reduction compared to males of the same age.

#### Retinal Nerve Fiber Layer (RNFL) thickness

The older age group demonstrated lower values, with an average of 1.27 µm in RNFL thickness compared to the younger age group (CI = [−1.51, −1.04] µm, p < 0.001). Female individuals showed thicker RNFL compared to males, with an average difference of 0.75 µm (CI = [0.50, 1.01] µm, p < 0.001). The interaction between sex and age group was non-significant (estimate = −0.25 µm, CI = [−0.58, 0.08] µm, p = 0.143), indicating that the age-related RNFL decline did not differ significantly between males and females.

#### Ganglion Cell-Inner Plexiform Layer (GC-IPL) thickness

The older age group showed lower values, with an average difference of 3.65 µm compared to the younger group (CI = [−4.12, −3.19] µm, p < 0.001). Female individuals had lower values compared to males, with an average difference of 1.10 µm (CI = [−1.38, −0.83] µm, p < 0.001). Additionally, a significant interaction between sex and age group was found, with females in the older age group showing on average 1.45 µm smaller values in GC-IPL thickness compared to males in the same group (CI = [1.04, 1.87] µm, p < 0.001), suggesting an attenuated age-related decline in GC-IPL thickness for females.

### Disease-related deviations from normative retinal decline

#### Macular thickness and subfields

As shown in Figure 2B, we found that age-related macular decline was generally more pronounced for mental disorders compared to the normative decline seen in controls, reflected by significantly negative deviations from the norm on average for macular thickness (z = −0.29, CI = [−0.35, −0.22], P < 0.0001). Sex-specific differences in these retinal thickness deviations did not reach statistical significance (all P > 0.3). The negative macular thickness deviations were driven by SZ (z = −0.47, CI = [−0.59, −0.35], P < 0.0001) and BD (z = −0.23, CI = [−0.33, −0.14], P < 0.0001), with a significant group difference between the two (Δz = 0.24, CI = [0.14, 0.33], P = 0.0001).

To investigate macular effects in more detail, we also looked at macular subfields (Figure 3A). Across mental disorders, we found that the age-related decline was generally more pronounced for the inner compared to the central (Δz_avg_ = −0.13, CI = [−0.06, −0.19], P = 0.0005; Figure 3B) and outer subfields (Δz_avg_ = −0.08, CI = [−0.02, −0.14], P = 0.01; Figure 3B). As shown in Figure 3C, disorder specific variability for SZ vs. BD was evident for the outer temporal subfield (Δz = −0.26, CI = [−0.41, −0.11], P = 0.0022) and the outer inferior subfield (Δz = −0.19, CI = [−0.35, −0.03], P = 0.0288); and for BD vs MDD for the inner inferior subfield (Δz = −0.21, CI = [−0.38, −0.03], P = 0.0253), inner nasal subfield (Δz = −0.21, CI = [−0.39, −0.03], P = 0.0288), and inner temporal subfield (Δz = −0.19, CI = [−0.36, −0.01], P = 0.0384).

**Figure 3.**
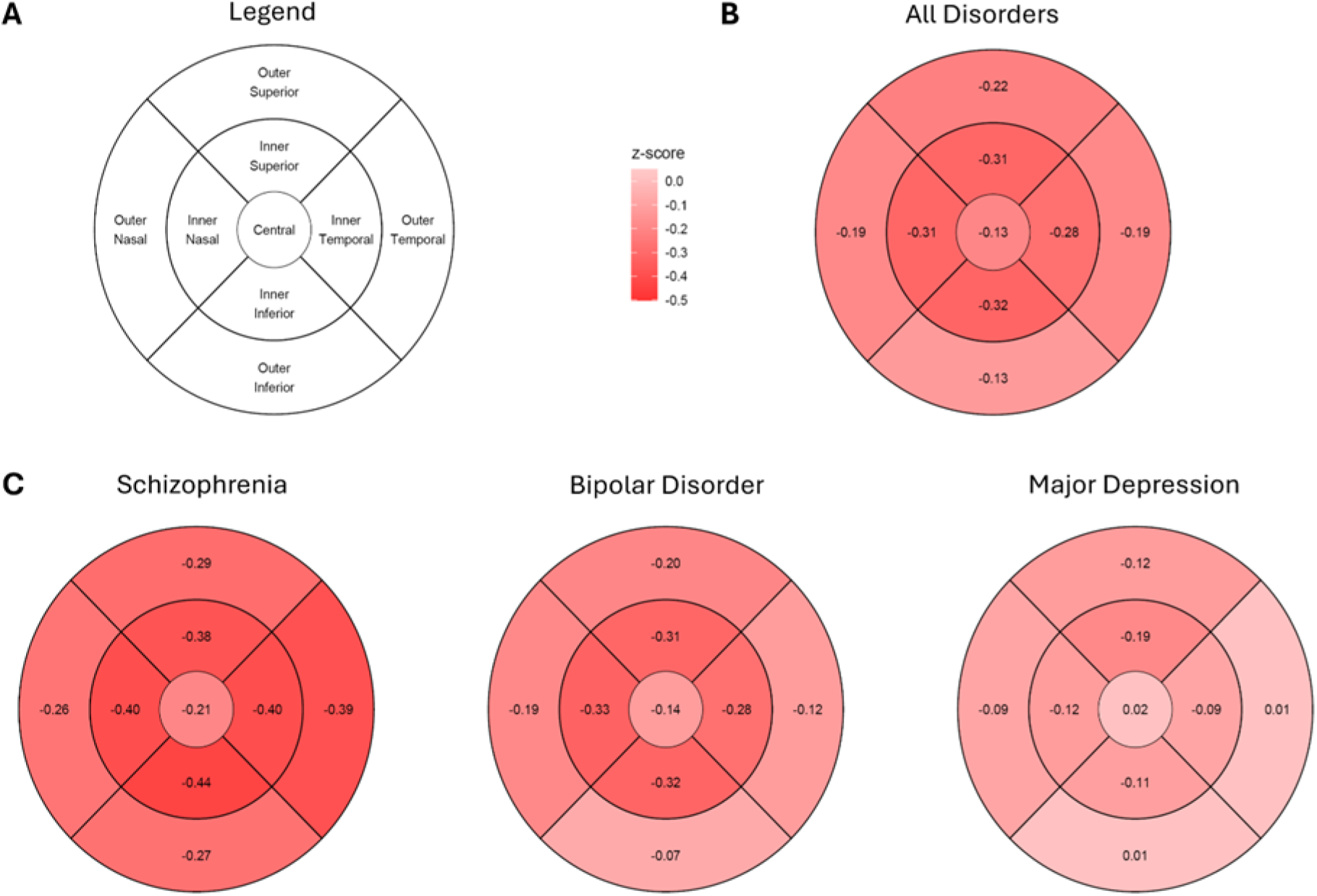
**A.** Schematic Representation of a Coronal Section of the macula, segregated in the subfields which are measured with OCT **B.** Average z-score differences of patients compared to HCs, more pronounced in the inner compared to the central and outer subfields **C.** Average z-score differences between patients of each disorder compared to HCs. The outer inferior and temporal subfields show significantly lower values in SZ compared to BD.

#### GC-IPL thickness

Age-related macular decline was also more pronounced across mental disorders for GC-IPL (z = −0.18, CI = [−0.25, −0.11], P < 0.0001), and again driven by SZ (z = −0.24, CI = [−0.36, −0.12], P = 0.001) and BD (z = −0.16, CI = [−0.26, −0.06], P = 0.004), with no significant difference between these two groups.

#### RNFL thickness

No evidence was found for pronounced RNFL decline across mental disorders (z = −0.05, CI = [−0.11, 0.01], P = 0.10) or individual diagnostic groups (all P > 0.09).

### Extreme value analysis

Our analysis of extreme negative z-scores (infranormal, i.e., below −1.96 SD) revealed significant enrichment of infranormal variability for macular thickness and GC-IPL thickness among participants with mental disorders, consistent with our analysis focused on average deviations (see above). Specifically, 4.65% of participants with mental disorders showed infranormal macular thickness values compared to 2.03% of healthy controls (X-squared = 17.65, df = 1, p = 0.0003), driven by several macular subfields (Figure S4). For GC-IPL, 4.83% showed infranormal values compared to 2.35% of controls (X-squared = 13.701, df = 1, p = 0.0002). Conversely, no such enrichment was found for RNFL thickness, with 1.07% of individuals with mental disorders compared to 1.80% of controls showing infranormal values, respectively (X-squared = 1.26, df = 1, p = 0.26). Finally, no evidence was found for enrichment of supranormal values for any retinal metric and diagnostic group (all p > 0.26).

### Robustness checks

#### Extended cardiovascular covariate analysis

Our extended model, i.e., using additional covariates that could have influenced retinal decline, confirmed the findings of our main analysis. Specifically, including body mass index, smoking status, alcohol drinker status, as well as accounting for a diagnosis of diabetes mellitus and socioeconomic status, we again found negative average deviations across mental disorders for macular thickness (z = −0.24, CI = [−0.32, −0.16], P < 0.0001) and GC-IPL (z = −0.18, CI = [−0.26, −0.10], P < 0.0001), but not RNFL (z = −0.01, CI = [−0.09, 0.07], P = 0.75). In line with our main analysis, no specific diagnostic group showed evidence for negative RNFL deviations (all P > 0.28). We also confirmed enrichment of infranormal values across mental disorders for macular thickness (X-squared = 18.35, df = 1, p < 0.0001), GC-IPL (X-squared = 10.18, df = 1, p = 0.0014) but not RNFL (X-squared = 2.01, df = 1, p = 0.156). Similarly, no evidence for enrichment of supranormal values was found (all p > 0.3).

#### Permutation analysis

Over 10 iterations of the permutation analysis, whereby in each iteration a random 80% sample of the HC sample was used for training, and all subsequent comparisons being made between the remaining 20% of HCs and patients, the average differences on the z-scores of the macular thickness was −0.28 (CI = [−0.37, −0.19], P < 0.0001), for the RNFL metric was 0.08 (CI = [−0.02, 0.19], P = 0.10), and for the GC-IPL was −0.16 (CI = [−0.26, −0.07], P = 0.0012). Similarly, regarding the effects observed within each individual disorder, the results of the permutation analysis showed high agreement with the original analysis.

## Discussion

Based on largescale data from 56,545 healthy UK Biobank individuals, this study provided a benchmark for comparing mental disorder-specific retinal decline to the normative age-related decline that one would expect in a healthy population. To this end, we estimated normative decline over the mid to late life span for several retinal parameters and showed that the retina declines curve-like and less pronounced in females compared to males. In a second step, we then used these retinal decline benchmarks for a comparison with the retinal decline seen in three major mental disorders (SZ, BD and MDD). We found that mental disorders generally deviated negatively from the norm, in line with the idea of an accelerated neuronal decline that is evident event in the most outward facing part of the central nervous system, which is the retina.

Our findings provide notable evidence of why variability should already be considered for healthy reference populations. The interactions of sex with age-related decline that we have found are a good example. As we’ve shown, while male individuals had higher macular and GC-IPL thickness in midlife, they demonstrated a steeper age-related thickness decline compared to female individuals, leading to a convergence of thickness measurements in later life. In contrast, RNFL thickness was on average higher in female individuals across the entire mid-to late lifespan, with age-related reductions following a similar trajectory in both sexes. While sex differences in metrics of retinal thickness have been shown before^38,40,41^, here we show that macular thickness trajectories across the lifespan are modulated by sex. These findings are consistent with the idea that neural structures of the retina reflect structural properties of the central nervous system. Indeed, while male individuals have been found to show higher absolute brain gray matter volumes in young and mid-adulthood, they also show a faster rate of decline^22,42^. Notably, both the absolute and the rate of change differences of these gray matter changes disappear once adjusting for total brain size^42^. Similarly, male individuals have higher axials lengths on average which could explain the differences we observe here since axial length has a known effect on retinal thickness measures^41^.

Once variability in healthy controls is taken into account and estimated with enough precision through normative modeling, it can be used as benchmark for the variability seen in mental disorders. Hence, by using our normative models as a reference against which we compared data from individuals with mental disorders, we revealed significant negative deviations from the normative population in total macular thickness and GC-IPL thickness but not RNFL thickness. This resulting pattern was also reflected in our extreme value analysis, where we revealed a marked enrichment of infranormal retinal thickness values in mental disorders compared to controls for macular thickness and GC-IPL. Intriguingly, these macular differences appeared to be more pronounced in the inner compared to the central and outer macular subfields. Combined with the negative deviations in GC-IPL, our findings could reflect the intrinsic neuroanatomy of the eye, with inner macular subfields having a higher density of ganglion cells and thus potentially being a better marker of the GC-IPL changes^43^. Interestingly, the inner subfields have also been shown to be more sensitive markers of sex-specific differences in macular thickness^38^. Together, these findings reinforce the utility of normative models in identifying specific patterns of retinal thinning associated with mental disorders and provide additional evidence for structural retinal abnormalities that may serve as peripheral indicators of neurodegeneration.

We then went on to explore the disease-specificity of these findings. Our direct comparison of the diagnostic groups revealed that individuals with SZ showed a more negative deviation in total macular thickness than individuals with BD, and that this effect was driven by the outer inferior and temporal macular subfields. While there is an abundance of case-control studies which have found thinner retinal layers in SZ and BD compared to healthy controls, their diagnostic and topological specificity has not been fully understood ^10,13,17^. Given the overlapping genetic and neurobiological underpinnings of SZ and BD ^44–48^, it is plausible that these disorders show both shared and distinct patterns of retinal degeneration. The more pronounced thinning in specific macular subfields in SZ could reflect disorder-specific neurodegenerative processes or differences in neuroinflammatory responses, aligning with findings of more severe cortical and subcortical structural anomalies in SZ. This suggests that retinal thickness deviations may serve as a peripheral marker of the differential neurodegenerative trajectories associated with each disorder.

Regarding macular thickness deficits in individuals with MDD, the current literature is equivocal^13^. The absence of such effects for individuals with MDD in our study adds to the evidence against a substantial macular decline in this diagnostic group. However, given the clinical heterogeneity of the phenotype, and the subtler structural brain alterations observed in MDD compared to SZ and BD ^2,5,19^, this question merits further study.

Biologically, the lack of significant findings in the macular RNFL – which consists of the axonal layer of retinal ganglion cells – may suggest that the observed thinning in the GC-IPL may stem from reductions in the cell bodies of retinal ganglion cells or a loss of surrounding extracellular matrix, rather than from changes in the axonal layer. Alternatively, the observed thinning might be localized within the inner plexiform layer, potentially reflecting reduced synaptic density in this region. This interpretation aligns with recent studies showing that the inner plexiform layer is particularly vulnerable to structural deficits in schizophrenia spectrum disorders^49^.

Taken together, the findings in our patient group partly confirm our hypothesis that the differences observed in neural structures of the retina would reflect brain structural differences of patients with mental disorders. SZ and BD patients showed indeed the most pronounced differences in the hypothesized order of magnitudes^19^. The absence of evidence for macular thickness differences in MDD patients in our study need not necessarily be interpreted as evidence for absence thereof. As in the case of the brain, these could be subtler and thus require a higher patient sample size to be estimated with precision ^19^.

There are two limitations that merit comment. First, because we relied on data from the UK Biobank, our normative models focused on the mid-to late lifespan, leaving early-life retinal changes outside the scope of this analysis. Although this limited the age range we could model, the large, standardized dataset with high quality phenotyping minimized measurement error and ensured consistency. Second, a more comprehensive understanding of age-related retinal decline would benefit from repeated measurements in the same individuals.

Our study also had several strengths. First, we leveraged the largest OCT dataset to date, which enabled us to establish a comprehensive normative benchmark and to compare multiple diagnostic groups both to this standard and to each other. Second, by using age- and sex-adjusted z-scores, we provided a statistically robust alternative to traditional case-control matching. Next, the extensive UK Biobank phenotyping allowed for adjustments for key variables, including OCT image quality, cardiovascular risk factors, socioeconomic status, and ethnicity. Finally, our comprehensive robustness checks confirmed the reliability and consistency of our findings across multiple conditions.

In conclusion, our study provided robust evidence of accelerated and disorder-specific retinal thinning in SZ and BD. These findings reinforce the retina’s potential as a sensitive, accessible marker of neurodegeneration in mental disorders and underscores the value of normative benchmarks for detecting both shared and distinct neurobiological changes across diagnostic groups.

## Data and Code availability

Data and Code availability All data analyzed in this study are available through the UK Biobank (http://www.ukbiobank.ac.uk/). All code will be made freely available after publication to ensure reproducibility at https://github.com/homanlab/.

## Data Availability

All data utilized in this study is publicly available at the UK Biobank (http://www.ukbiobank.ac.uk/). All code will be made freely available after publication to ensure reproducibility at https://github.com/homanlab

## Supplementary Results

### Accounting for eye laterality

Repeating our main analysis (Disease-related deviations from normative retinal thickness) using a mixed model for repeated measures (left and right eyes) instead of averaging the z-scores of the two eyes, we obtain highly similar the same results. We find covariate-adjusted deviations across diagnostic groups in macular thickness (z = −0.29, CI = [−0.35, −0.22], P < 0.001) and GC-IPL (z = −0.19, CI = [−0.25, −0.12], P < 0.001), but not RNFL (z = −0.05, CI = [−0.11, 0.01], P = 0.09). Sex-specific vulnerabilities to these retinal thickness deviations did not reach statistical significance (all P > 0.3). MT deviations were driven by SZ (z = −0.47, CI = [−0.59, −0.35], P < 0.001) and BD (z = −0.23, CI = [−0.33, −0.14], P < 0.001), with a significant group difference between the two (Δz = 0.24, CI = [0.14, 0.33], P < 0.001). Similarly, GC-IPL deviations were driven by SZ (z = −0.25, CI = [−0.37, −0.13], P < 0.001) and BD (z = −0.16, CI = [−0.26, −0.06], P = 0.003). In line with the patient group analysis, no specific patient group showed significant RNFL deviations (all P > 0.09). MDD patients showed no significant deviations in any of the analyzed retinal metrics (all P > 0.07). No significant differences were observed between female and male patients (all P > 0.4).

### Interaction effects in normative retinal trajectories

Our extensive mixed modeling on the healthy control (HC) individuals which aimed to describe the normative curves of macular thickness, retinal nerve fibre layer (RNFL) and ganglion cell-inner plexiform layer (GC-IPL) showed that the laterality effects observed in the simpler models, with the right eye showing on average higher thickness across all 3 metrics (see also main Results section Normative Trajectories of retinal thickness), interacted significantly with age and sex in macular thickness and RNFL, but not GC-IPL. Specifically, as outlined below the left-right discrepancy was generally higher in females and the older age group (age >65 years). In all models described below, the reference group are male individuals, left-eye measurements and age <45 years.

#### Total macular thickness

For macular thickness, the interaction between sex and side indicated that the difference between the right and left eyes was more pronounced in females. Female individuals exhibited an additional 1.06 µm (95% CI: 0.71 to 1.41 µm, *p* < 0.001) greater macular thickness in the right eye compared to males, suggesting that the laterality effect is more pronounced in females.

A significant interaction effect was additionally observed between laterality and age group, with the right macular thickness of participant in the right age (65+) 0.46 µm (95% CI: 0.11 to 0.82 µm, *p* = 0.011) slightly higher macular thickness in the right eye compared to younger participants, with.

#### Retinal nerve fiber layer thickness

For RNFL, the interaction between sex and laterality indicated that females had a more pronounced difference thickness difference between the right and left eyes than males, with the RNFL being 0.87 µm thicker on the right side (95% CI: 0.66 to 1.07 µm, *p* < 0.001). The interaction between age group and side suggested that older participants (65+) had a slightly greater difference between right and left eyes compared to younger participants, with a 0.30 µm increase (95% CI: 0.10 to 0.51 µm, *p* = 0.004).

Ganglion cell-inner plexiform layer thickness.

For GC-IPL, the interaction between sex and laterality was not statistically significant (0.10 µm, 95% CI: −0.05 to 0.25 µm, *p* = 0.179), indicating no strong laterality differences between males and females. Similarly, the interaction between age group and side was not significant (estimate = 0.09 µm, 95% CI: −0.06 to 0.24 µm, *p* = 0.226).

### Robustness analyses

#### Patients with prior depressive symptoms

Using a more permissive definition of depressive symptoms, i.e. patients with a lifetime history of depressive symptoms lasting more than one week (instead of the F33 ICD code), we found again, no negative thickness deviations in any of the retinal metrics. Reflecting the high prevalence of depressive symptoms, 29,025 of the original 56,545 HC participants with complete OCT and covariate data belonged to the depressive symptom group. We followed the same analytical pipeline outlined in the main Methods section for our normative model analysis and the estimation of the z-scores. The 27,520 HCs who did not report such depressive symptoms were used as the normative cohort.

In alignment with our original analysis, in all 3 retinal metrics, patients with depressive symptoms showed no negative thickness deviations, but rather statistically significant yet marginal in terms of Δ *positive* retinal thickness deviations in MT (Δz = 0.02, CI = [0.00, 0.03], P = 0.024), GC-IPL (z = 0.02, CI = [0.01, 0.04], P = 0.005), and RNFL (Δz = 0.04, CI = [0.03, 0.06], P < 0.0001). Meanwhile, compared to this smaller HC group based on which the normative model was trained, the negative thickness deviations of SZ and BD stayed consistent with our original results. For SZ there were significant negative deviations in MT (Δz = −0.47, CI = [−0.61, −0.33], P < 0.0001), GC-IPL (Δz = −0.23, CI = [−0.37, −0.09], P = 0.001), but not RNFL (Δz = −0.06, CI = [−0.19, 0.06], P = 0.303). For BD there were significant negative deviations in MT (Δz = −0.22, CI = [−0.33, −0.12], P = 0.0001), GC-IPL (Δz = −0.16, CI = [−0.27, −0.05], P = 0.008), but not RNFL (Δz = −0.06, CI = [−0.16, 0.04], P = 0.248).

#### Direct comparison of HC and patient group using linear modeling

Averaging the results of 250 permutations, which compared 1614 resampled HC to the patient group (N = 538) we observed significant reductions in macular thickness, and ganglion cell-inner plexiform layer (GC-IPL) thickness and non-significant reductions in retinal nerve fiber layer (RNFL) thickness across diagnostic groups. The magnitudes we observed are well in alignment with our normative model analysis.

#### Macular thickness

For the combined patient group, a moderate effect size was observed, with a mean Cohen’s D of −0.25 (−3.93 μm, P < 0.0001). The SZ group exhibited the greatest effect, with a mean Cohen’s D of −0.41 (−6.97 μm, Pavg < 0.0001), followed by the BD group, with a mean Cohen’s D of −0.20 (−3.00 μm, Pavg = 0.0025). The effect for the MDD group was consistently non-significant with a mean Cohen’s D of −0.08 (−1.11 μm, Pavg = 0.455).

#### Retinal nerve fiber layer

The combined patient group, a small effect size showed an average non-significant difference from the HC (Cohen’s D = −0.04, −0.32 μm, Pavg = 0.237). The same was true across the diagnostic subgroups; SZ: (Cohen’s D= −0.10, −0.44 μm, Pavg = 0.275). BD: (Cohen’s D = - 0.07 (−0.47 μm, P = 0.161). MDD (Cohen’s D = +0.12, +0.30 μm, Pavg = 0.545).

#### Ganglion Cell-Inner Plexiform Layer (GC-IPL)

For the combined patient group, a small to moderate effect size was observed, with a mean Cohen’s D of −0.14 (−1.02 μm, P = 0.0026). The SZ group exhibited the greatest effect: mean Cohen’s D of −0.19 (−1.38 μm, P = 0.0069), followed by BD: mean Cohen’s D of −0.12 (−0.89 μm, P = 0.0407). The effect for MDD was non-significant: mean Cohen’s D of −0.08 (−0.76 μm, P = 0.238).

Overall, these results are very consistent with our original normative modeling analysis.

### Extended cardiovascular covariate analysis

In addition to our original covariates, including body mass index, smoking status, alcohol drinker status, socioeconomic status, as well as accounting for a diagnosis of diabetes melitus, we observed covariate-adjusted deviations across diagnostic groups in MT (z = −0.24, CI = [−0.32, −0.16], P < 0.0001) and GC-IPL (z = −0.18, CI = [−0.26, −0.10], P < 0.0001), but not RNFL (z = −0.01, CI = [−0.09, 0.07], P = 0.75).

MT deviations were driven by SZ (z = −0.40, CI = [−0.54, −0.26], P < 0.0001) and BD (z = - 0.19, CI = [−0.28, −0.10], P = 0.0011), with a significant group difference between the two (Δz = 0.21, CI = [0.12, 0.30], P = 0.001). MDD patients showed no significant deviations in MT (z = −0.09, CI = [−0.27, 0.09], P = 0.36).

Similarly, GC-IPL deviations were driven by SZ (z = −0.22, CI = [−0.37, −0.07], P = 0.0024) and BD (z = −0.16, CI = [−0.27, −0.05], P = 0.0057). MDD patients showed no significant deviations in GC-IPL (z = −0.15, CI = [−0.33, 0.03], P = 0.11).

In line with our main analysis, no specific patient group showed significant RNFL deviations (all P > 0.28). MDD patients showed no significant deviations in any of the analyzed retinal metrics (all P > 0.11). No significant differences were observed between female and male patients (all P > 0.4).

### Outlier removal-IQR multiplier robustness analysis

To show that the choice of IQR multiplier for outlier removal does not significantly alter our results we repeated our analysis across a range of IQR values, from 1 to 7, in increments of 0.5. As shown in Figure S1, the effect sizes and significance of our results do not change across this wide range of IQRs, with the exception of a significant reduction in the RNFL of BD patients at IQRs lower than 3.

---

**Figure S1.**
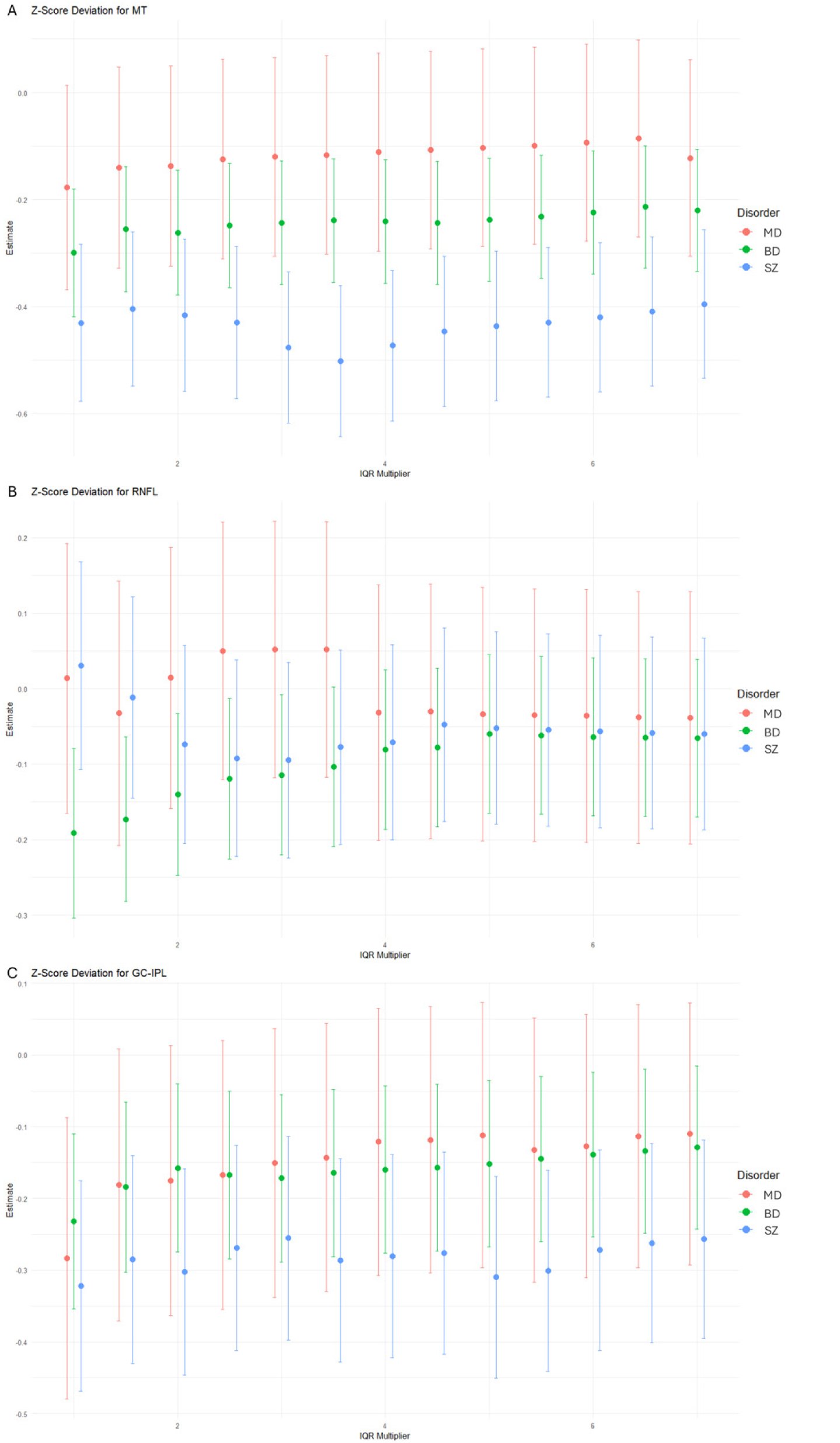
Plotting of disorder-specific average deviation scores across IQR multipliers for (A) macular thickness (B) RNFL and (C) GC-IPL.

**Figure S2.**
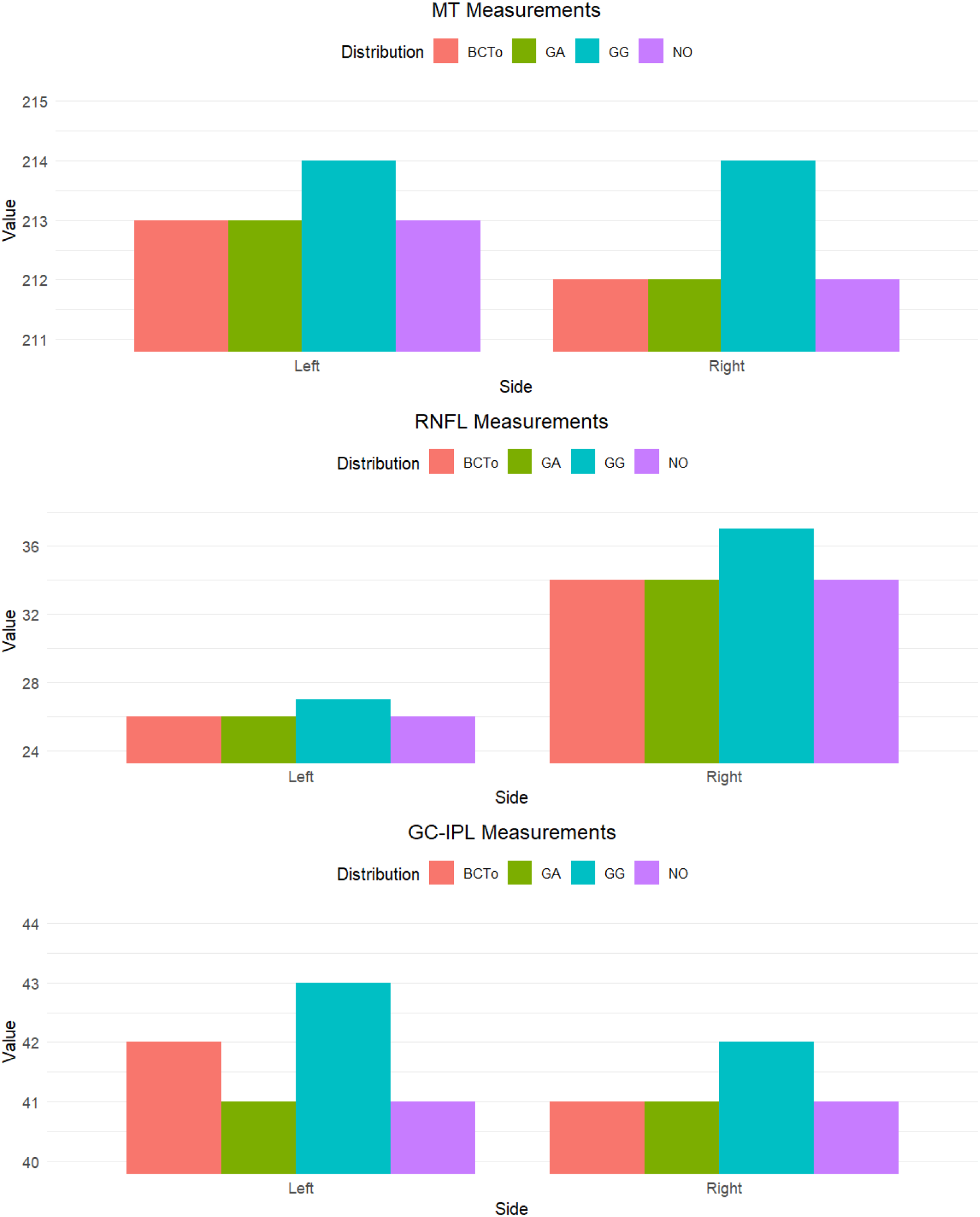
The Mean Squared Errors (MSE) of 5-fold 80-20 cross-validation (CV) of 5 distribution fits for the GAMLSS model were compared: the Box-Cox Transformed (BCTo), Gamma (GA), Generalized Gamma (GG) and normal (NO). The Normal Distribution (NO) in our study shows consistently equivalent performance to more complex distributions such as the Generalized Gamma or the BCTo in terms of MSE. Based on these empirical findings and since it represents the more parsimonious model, we fit our main *gamlss* model with the normal distribution.

**Figure S3.**
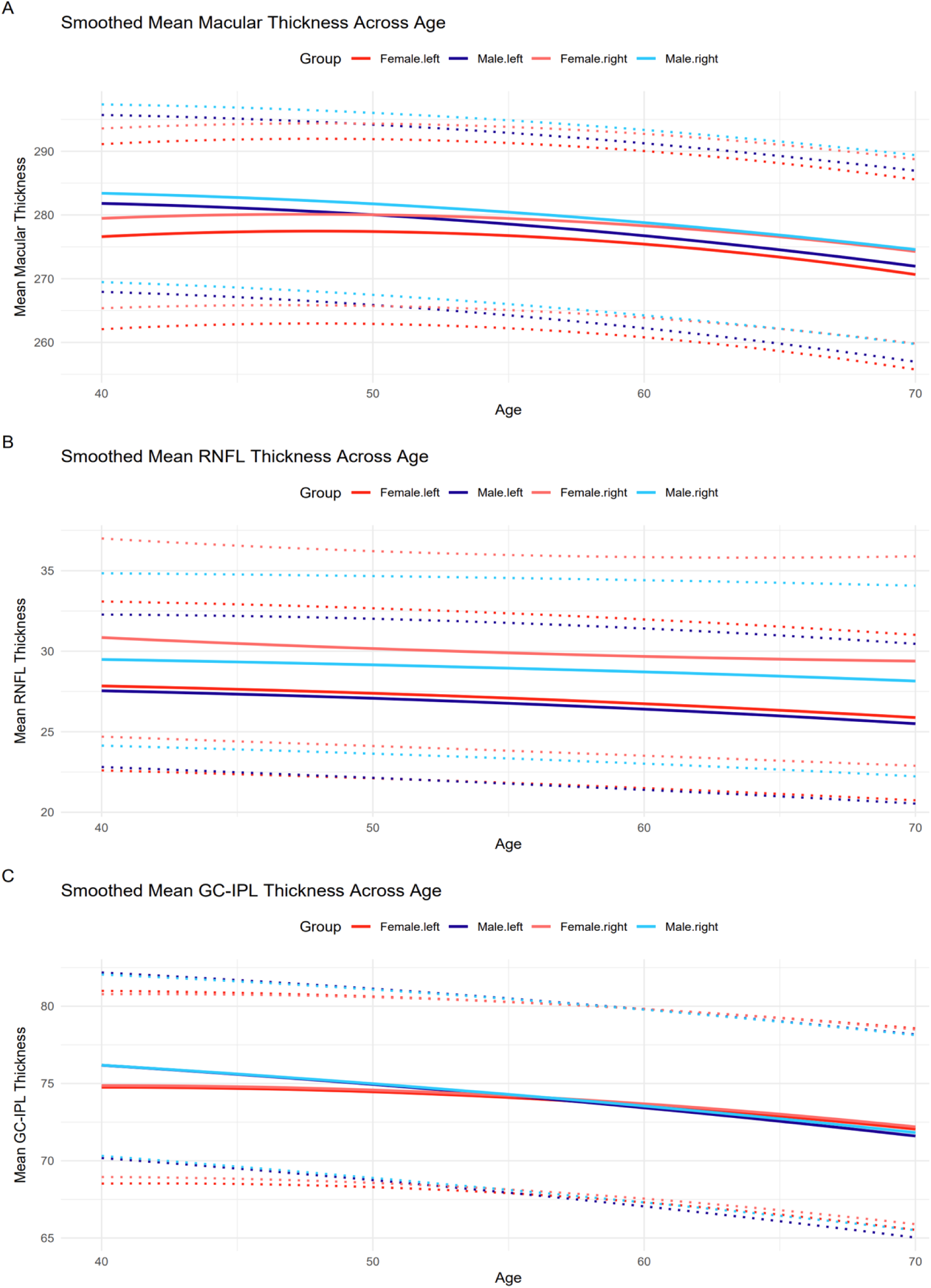
Plotting of lateralized normative models of retinal thickness across age. The Y-axis values represent the predicted normative values for the covariate sample modes (image quality score between 63 and 65 and reported British ethnicity). Left and right sides show very good convergence, justifying the averaging of the values/ z-scores into one value to simplify downstream analyses.

**Figure S4.**
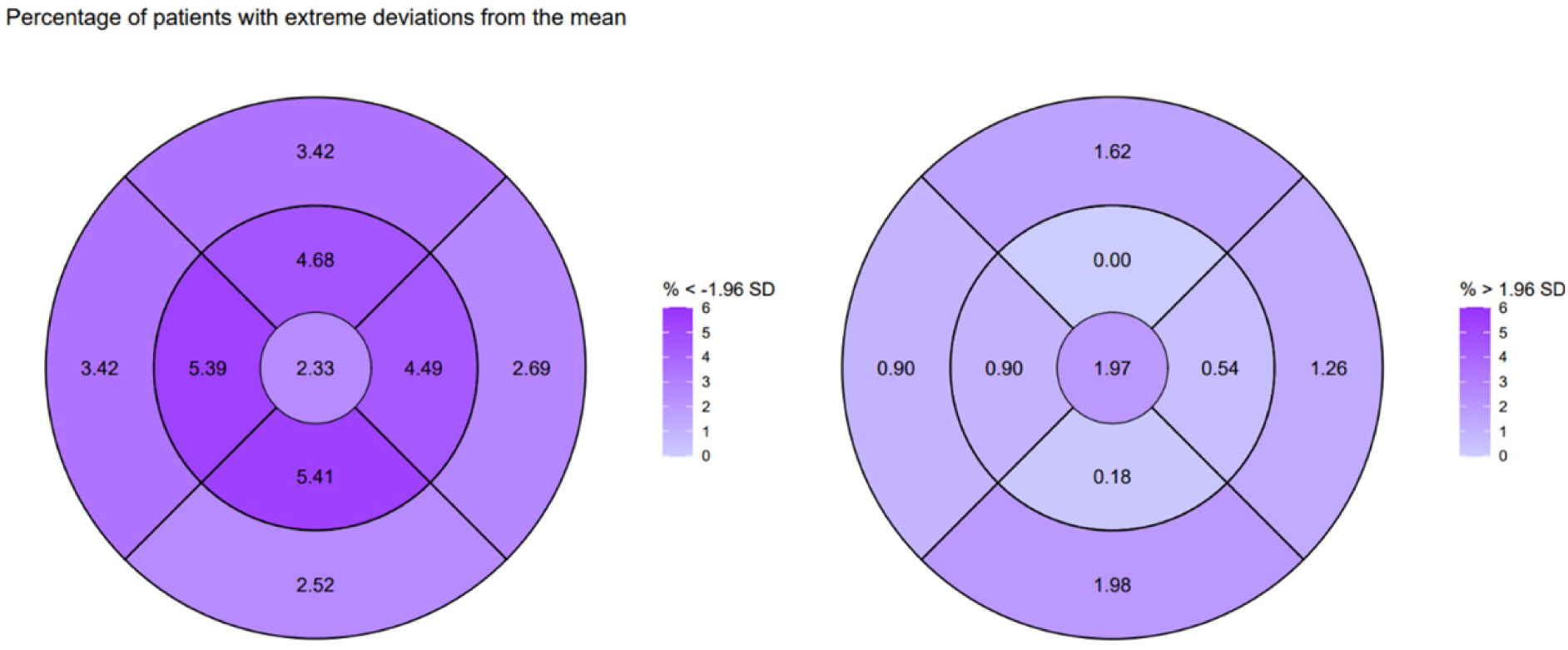
Percentage of individuals from the combined patient group with “extreme” low (left) and high (right) subfield thickness; based on the normal distribution we expect 2.5% of individuals symmetrically in each tail. On the left we observe an increase (>2.5%) of individuals with infranormal thickness and on the right a reduction (<2.5%) of individuals with supranormal thickness.

